# Time trends in socio-economic inequalities in HIV testing: insights from population-based surveys in 16 sub-Saharan African countries

**DOI:** 10.1101/19006015

**Authors:** Pearl Anne Ante-Testard, Tarik Benmarhnia, Anne Bekelynck, Rachel Baggaley, Eric Ouattara, Laura Temime, Kévin Jean

## Abstract

**Background:** Overall increase in the uptake of HIV testing in the past decades may hide discrepancies across socio-economic groups. We used population-based surveys conducted in sub-Saharan Africa to quantify socio-economic inequalities in recent HIV testing uptake, together with their trends over the two past decades.

**Methods:** We analyzed the data from Demographic and Health Surveys in sub-Saharan African countries where at least one survey was conducted before and after 2008. Country- and gender-specific proportions of recent (<12 month) HIV testing were assessed across wealth and education groups, and inequalities were quantified using the relative and slope indices of inequalities. Time trends in inequalities were assessed and results were pooled across countries using random-effect meta-analyses.

**Findings:** We analyzed data from 32 surveys conducted between 2003 and 2016 in 16 countries among 537,784 participants. In pre-2008 surveys, women reported higher HIV testing uptake than men in 8 out of 16 countries, and in 15 out of 16 countries in post-2008 surveys. After 2008, the wealthiest women were on average 2.77 (95% CI 1.42-5.40) times more likely to report recent testing than the poorest; and 3.55 (1.85-6.81) times in men. The averaged absolute difference in recent testing between the richest and poorest was 11.1 (4.6-17.5) percentage points in women and 15.1 (9.6-20.6) in men. Over time, relative inequalities in recent HIV testing decreased in both genders, while absolute inequalities plateaued in women and increased in men.

**Interpretations:** The overall increase in HIV testing uptake that was stimulated by the impetus to scale up HIV treatment in sub-Saharan Africa led to a decrease in relative inequalities, while absolute inequalities persisted. Within most countries, large inequalities still remained, both in absolute and relative scales, especially in West and Central Africa. A greater focus should be put on equity in monitoring HIV testing programs.

A French version of this article is available in the Appendices [Une version française de cet article est disponible en appendice].

**Funding:** INSERM-ANRS (France Recherche Nord & Sud Sida-HIV Hépatites), grant number ANRS-12377.

## Introduction

As the gateway to many HIV prevention and care services, including antiretroviral therapy (ART), HIV testing has played a central role in the HIV response. As ART became increasingly available in most countries, HIV testing strategies have evolved over time, from a cautious approach which focused on counselling and confidentiality, to a push to increase routine access to testing in clinical settings and through community approaches at large scale.^1,2^ This translated into a significant increase in the access to and uptake of HIV testing in many countries.

In Africa in particular, the percentage of people living with HIV (PLHIV) knowing their status increased from 10% in 2005 to 85% in eastern and southern Africa and 64% in western and central Africa in 2018.^3^ However, an estimated 1.1. million PLHIV in eastern and southern Africa 1.3 million in western and central Africa remain unaware of their status. This means, efforts are still needed to reach the target of 90% of PLHIV knowing their status by 2020, which is the first 90 of the global 90-90-90 target adopted by the Joint UN Programme on HIV/AIDS (UNAIDS).^4^ It is also essential to ensure that no specific group of the population are left behind in the way toward these objectives.

Several cross-sectional studies conducted in sub-Saharan Africa have reported lower uptake of HIV testing in the poorest or less-educated population groups.^5–9^ Whether these inequalities increased or decreased during the intensification of HIV testing activities is still unknown. Indeed, the scale-up of health interventions does not necessarily translate into reduced health inequalities, and may even exacerbate these inequalities. For instance, programs that increased cancer screening services were recently shown to have failed in reducing socio-economic inequalities in uptakes.^10,11^ Monitoring time trends in socio-economic inequalities in response to expanded HIV testing is thus essential to assess and ensure equity of HIV programs in line with the Sustainable Development Goals.

In this study, based on population-based surveys conducted in several sub-Saharan African countries, we assess time trends in both relative and absolute socio-economic inequalities in the uptake of HIV testing during the time of HIV testing progression and ART scale-up.

## Material and methods

We analysed data collected from the Demographic and Health Surveys (DHS). DHS are nationally-representative cross-sectional surveys, collecting data on a wide range of health indicators, including specific data on HIV/AIDS. Based on a multistage design with households as sampling units, all adults (generally aged 15-49 years) from selected households are eligible. Depending on the survey, data among men and/or HIV indicators may be collected among a sub-sample of the selected households only. Consenting adults are interviewed face-to-face by trained interviewers using a standardized questionnaire. Questionnaire items cover various dimensions such as socio-demographic characteristics, sexual behaviours, reproductive health as well as a specific section focusing on HIV-related issues.^12^ Data from the DHS surveys are publicly available for academic research (https://dhsprogram.com/).

We selected sub-Saharan African countries in which at least one DHS survey covering HIV indicators has been conducted both before and after 2008, i.e. before and after the release of international recommendations to expand provider-initiated opt-out testing.^13^ With the recommendation of provider-initiated testing, the profile of HIV-testing users may have changed from a small self-selecting group to a wider part of the population accessing health services.^5^ For countries where multiple surveys were available in one of the time period, we only considered the most recent one (as available up to March 2019). Pre- and post-2008 surveys were thereafter termed earlier and later surveys, respectively.

### Data collected

Each household was classified as rural or urban based on nationally-defined boundaries. Individual socio-demographic characteristics collected as part of the interview included age, educational level (none, primary, secondary/higher) and family status. Household wealth was assessed through the DHS’ wealth index, a composite measure of living standard based on a set of the household assets (e.g. television, refrigerator) and characteristics (e.g. type of water access, type of flooring).^14^

Participants were asked whether they had ever been tested for HIV, and if so, the time since their last test. The outcome of analysis was the self-report of having completed a recent (<12 months) HIV test.

### Statistical analysis

Firstly, for each survey, the proportion of participants reporting recent HIV testing was calculated overall and per wealth quintile, while accounting for survey design and sampling weights.

Secondly, for each survey round, we assessed within-country inequalities based on participants’ relative rank in the cumulative distribution of the wealth index. Inequalities were then measured both on relative and absolute scales. Reporting inequalities both in relative and absolute terms is highly recommended, especially when monitoring changes, as conclusions may diverge when based only on one or the other.^15^ We used the relative index of inequality (RII) and the slope index of inequality (SII) to measure relative and absolute inequalities, respectively.^16,17^ The RII expresses the ratio of the predicted outcome between the richest and the poorest of the wealth distribution, while the SII represents the absolute difference in the predicted proportions of these two extremes. Both indicators were obtained using a modified Poisson regression with robust variance and a log link function to estimate the association between participants’ relative wealth rank and recent HIV testing, and using generalized estimating equations to account for the clustering of observations.^18,19^

Thirdly, we assessed time trends in relative and absolute inequalities in the recent HIV testing. For each country, we computed the ratio of RIIs between the later and the earlier surveys, *RII ratio* = *RII* _*post*−2008/_*RII*_*pre*−2008_, as well as the difference of SIIs : *SII difference* = *SII* _*post*−2008_ − *SII* _*pre*−2008_, together with their 95% confidence interval (CI). These trends indicators were standardized on the number of years elapsed between the earlier and the later surveys (Appendix 1). A RII-ratio value >1 (respectively <1) thus reflects increasing (respectively decreasing) relative inequalities, while a SII-difference >0 (respectively <0) reflects increasing (respectively decreasing) absolute inequalities.

Lastly, we pooled inequalities estimates across countries for each survey round, as well as trends indicators, using random-effects meta-analyses.^20^ Between-countries heterogeneity was assessed using *I*^2^ statistics. Inequalities may differ according to the dimension considered for their measure. We thus reproduced all analyses using the relative rank in the cumulative distribution of educational attainment instead of wealth. As attitudes toward testing are likely to depend on gender, all analyses were stratified according to sex. All analyses were conducted using the R software version 3.6.0.

## Results

### Study population

Sixteen sub-Saharan African countries were included in the analyses, namely: Sierra Leone, Guinea, Liberia, Côte d’Ivoire, Mali, Niger, Cameroon, the Democratic Republic of Congo (Congo DR), Zambia, Lesotho, Zimbabwe, Rwanda, Malawi, Tanzania, Kenya and Ethiopia. The earlier surveys were conducted between 2003 and 2008 and the later surveys between 2008-09 and 2016, with an inter-survey time period ranging from 5 to 11 years across countries (Table 1).

**Table 1:** Survey and population characteristics, by country, survey round and gender. Countries are ordered west to east. F: Female; M: Male.

| *West-Central Africa* |  | Sierra Leone |  |  |  | Guinea |  |  |  | Liberia |  |  |  | Côte d'Ivoire |  |  |  |
| --- | --- | --- | --- | --- | --- | --- | --- | --- | --- | --- | --- | --- | --- | --- | --- | --- | --- |
| Survey Year |  | 2008 |  | 2013 |  | 2005 |  | 2012 |  | 2007 |  | 2013 |  | 2005 |  | 2011-12 |  |
| Gender |  | F | M | F | M | F | M | F | M | F | M | F | M | F | M | F | M |
| *\*Individual response rate (%)* |  | 94 | 93 | 97 | 96 | 97 | 95 | 98 | 97 | 95 | 93 | 98 | 95 | 90 | 88 | 93 | 91 |
| Sample size (n) |  | 7,374 | 3,280 | 16,658 | 7,262 | 7,954 | 3,174 | 9,142 | 3,782 | 7,092 | 6,009 | 9,239 | 4,118 | 5,183 | 4,503 | 10,060 | 5,135 |
| Living in rural area (%) |  | 64.0 | 62.8 | 64.4 | 62.8 | 68.9 | 61.4 | 63.7 | 60.6 | 57.7 | 59.6 | 39.0 | 41.4 | 52.7 | 53.0 | 48.6 | 49.7 |
| Age (%): 15-24 years |  | 32.3 | 28.3 | 39.4 | 34.2 | 35.2 | 36.1 | 40 | 35.9 | 37.7 | 36.5 | 40.3 | 38.5 | 45.5 | 40.8 | 39.5 | 33.9 |
| 25-34 years |  | 36.4 | 25.0 | 30.8 | 25.1 | 29.9 | 19.5 | 30.4 | 24.3 | 29.9 | 28.0 | 30.4 | 30.3 | 30.8 | 31.7 | 34.1 | 29.4 |
| ≥ 35 years |  | 31.2 | 46.7 | 29.8 | 40.8 | 34.9 | 44.4 | 29.5 | 39.8 | 32.4 | 35.5 | 29.3 | 31.2 | 23.7 | 27.5 | 26.4 | 36.8 |
| Family situation: Living in union |  | 74.9 | 63.3 | 65.5 | 57.1 | 79.1 | 59.2 | 73.6 | 54.9 | 64.0 | 56.8 | 58.3 | 53.9 | 59.0 | 44.4 | 62.7 | 52.7 |
| Single |  | 19.0 | 33.3 | 28.4 | 39.3 | 16.5 | 36.6 | 22.5 | 43.3 | 26.1 | 37.9 | 31.0 | 42.5 | 32.3 | 49.7 | 30.2 | 42.4 |
| Widowed/ separated |  | 6.1 | 3.4 | 6.2 | 3.5 | 4.4 | 4.2 | 4.0 | 1.7 | 9.9 | 5.3 | 10.7 | 3.7 | 8.7 | 5.9 | 7.1 | 4.9 |
| Educational level (%): None |  | 65.9 | 50.1 | 55.8 | 42.8 | 77.5 | 51.2 | 67 | 42.9 | 42.4 | 17.6 | 33.2 | 12.9 | 53.9 | 34.0 | 53.2 | 35.9 |
| Primary |  | 13.0 | 13.8 | 14.0 | 12.2 | 11.4 | 16.7 | 13.9 | 17.9 | 32.9 | 33.3 | 31.1 | 29.2 | 26.5 | 25.1 | 25.4 | 26.3 |
| Secondary |  | 21.1 | 36.1 | 30.2 | 45.0 | 11.1 | 32.1 | 19.1 | 39.1 | 24.6 | 49.1 | 35.7 | 57.9 | 19.6 | 40.9 | 21.4 | 37.8 |
| HIV prevalence (%) |  | 1.7 | 1.1 | 1.7 | 1.2 | 1.9 | 1.1 | 2.1 | 1.4 | 1.9 | 1.2 | 2.4 | 1.8 | 6.2 | 2.8 | 4.6 | 3.3 |
| Recent uptake of HIV testing (%) |  | 5.3 | 4.8 | 17.6 | 8.2 | 1.3 | 3.3 | 4.9 | 5.1 | 2.0 | 2.8 | 21.6 | 13.7 | 4.8 | 4.4 | 15.4 | 9.9 |
|  |  | Mali |  |  |  | Niger |  |  |  | Cameroon |  |  |  | Congo DR |  |  |  |
| Survey Year |  | 2006 |  | 2012-13 |  | 2006 |  | 2012 |  | 2004 |  | 2011 |  | 2007 |  | 2013-14 |  |
| Gender |  | F | M | F | M | F | M | F | M | F | M | F | M | F | M | F | M |
| *\*Individual response rate (%)* |  | 97 | 91 | 96 | 93 | 96 | 92 | 95 | 88 | 94 | 93 | 97 | 96 | 97 | 95 | 99 | 97 |
| Sample size (n) |  | 14,583 | 4,207 | 10,424 | 4,399 | 9,223 | 3,549 | 11,160 | 3,928 | 10,656 | 5,280 | 15,426 | 7,191 | 9,995 | 4,757 | 18,827 | 8,656 |
| Living in rural area (%) |  | 66.3 | 63.7 | 75.2 | 74.9 | 80.3 | 74.4 | 81.2 | 75.4 | 45.2 | 42.7 | 46.5 | 45.0 | 54.6 | 57.0 | 61.6 | 63.0 |
| Age (%): 15-24 years |  | 39.6 | 35.9 | 35.8 | 29.1 | 36.5 | 31.3 | 34.2 | 28.0 | 46.3 | 41.2 | 43.2 | 39.2 | 43.1 | 39.1 | 41.2 | 36.4 |
| 25-34 years |  | 31.5 | 23.1 | 35.9 | 24.3 | 34.1 | 25.3 | 37.0 | 24.8 | 29.1 | 27.1 | 29.7 | 26.4 | 30.0 | 25.6 | 32.7 | 26.4 |
| ≥ 35 years |  | 28.8 | 41.0 | 28.2 | 46.7 | 29.4 | 43.5 | 28.8 | 47.2 | 24.6 | 31.6 | 27.0 | 34.4 | 26.9 | 35.3 | 26.1 | 37.2 |
| Family situation: Living in union |  | 84.8 | 65.1 | 84.6 | 67.6 | 86.1 | 66.5 | 88.5 | 69.8 | 67.2 | 50.7 | 62.9 | 50.3 | 66.3 | 56.5 | 64.2 | 58.2 |
| Single |  | 11.8 | 31.2 | 13.6 | 31.6 | 9.9 | 31.3 | 7.9 | 28.6 | 24.0 | 40.1 | 28.3 | 45.0 | 24.3 | 38.3 | 26.0 | 37.5 |
| Widowed/ separated |  | 3.4 | 3.7 | 1.8 | 0.8 | 4.0 | 2.2 | 3.6 | 1.5 | 8.7 | 9.2 | 8.8 | 4.8 | 9.4 | 5.2 | 9.7 | 4.2 |
| Educational level (%): None |  | 78.2 | 60.2 | 75.8 | 62.0 | 83.5 | 68.5 | 80.1 | 62.9 | 22.4 | 11.5 | 20.9 | 9.5 | 20.8 | 6.3 | 15.4 | 4.1 |
| Primary |  | 11.4 | 19.3 | 9.3 | 13.6 | 10.4 | 17.3 | 11.4 | 19.3 | 38.6 | 36.7 | 32.9 | 33.2 | 38.5 | 29.9 | 36.9 | 22.3 |
| Secondary |  | 10.3 | 20.5 | 14.9 | 24.4 | 6.1 | 14.1 | 8.5 | 17.7 | 39.1 | 51.8 | 46.1 | 57.3 | 40.6 | 63.8 | 47.7 | 73.6 |
| HIV prevalence (%) |  | 1.5 | 1.1 | 1.3 | 0.9 | 0.7 | 0.7 | 0.3 | 0.4 | 6.6 | 3.9 | 5.6 | 2.9 | 1.6 | 0.9 | 1.6 | 0.6 |
| Recent uptake of HIV testing (%) |  | 3.4 | 3.5 | 6.9 | 6.5 | 1.0 | 2.1 | 8.4 | 2.7 | 5.3 | 7.8 | 23.6 | 21.4 | 4.7 | 4.5 | 9.1 | 7.6 |

\*Based on each country's USAID DHS Final Report.

| *Eastern-Southern Africa* |  | Zambia |  |  |  | Lesotho |  |  |  | Zimbabwe |  |  |  | Rwanda |  |  |  |
| --- | --- | --- | --- | --- | --- | --- | --- | --- | --- | --- | --- | --- | --- | --- | --- | --- | --- |
| Survey Year |  | 2007 |  | 2013-14 |  | 2004 |  | 2014 |  | 2005-06 |  | 2015 |  | 2005 |  | 2014-15 |  |
| Gender |  | F | M | F | M | F | M | F | M | F | M | F | M | F | M | F | M |
| *Individual response rate (%) |  | 97 | 91 | 96 | 91 | 94 | 85 | 97 | 94 | 90 | 82 | 96 | 92 | 98 | 97 | 99.5 | 99.5 |
| Sample size (n) |  | 7,146 | 6,500 | 16,411 | 14,773 | 7,095 | 2,797 | 6,621 | 2,931 | 8,907 | 7,175 | 9,955 | 8,396 | 11,321 | 4,820 | 13,497 | 6,217 |
| Living in rural area (%) |  | 57.9 | 56.8 | 53.8 | 53.9 | 76.3 | 78.5 | 63.5 | 66.2 | 60.7 | 59.5 | 61.5 | 64.0 | 83.0 | 82.6 | 80.5 | 80.0 |
| Age (%): |  |  |  |  |  |  |  |  |  |  |  |  |  |  |  |  |  |
|  | 15-24 years | 41.2 | 38.2 | 40.4 | 38.4 | 44.7 | 44.7 | 41.8 | 42.7 | 46.1 | 46.8 | 39.1 | 41.2 | 43.6 | 42.5 | 38.7 | 36.6 |
|  | 25-34 years | 33.9 | 29.7 | 32.2 | 26.2 | 26.2 | 24.3 | 31.0 | 25.4 | 30.1 | 27.4 | 32.9 | 27.0 | 28.3 | 23.7 | 33.0 | 30.2 |
|  | ≥ 35 years | 25.0 | 32.1 | 27.4 | 35.4 | 29.1 | 31.0 | 27.3 | 31.9 | 23.8 | 25.8 | 28 | 31.8 | 28.1 | 33.9 | 28.3 | 33.2 |
| Family situation: Living in union |  | 61.6 | 55.8 | 60.1 | 55.1 | 52.3 | 42.6 | 54.6 | 40.0 | 57.7 | 47.7 | 61.8 | 51.5 | 48.7 | 51.9 | 51.7 | 54.2 |
|  | Single | 26.0 | 39.3 | 27.9 | 40.6 | 33.4 | 50.8 | 33.1 | 51.5 | 27.0 | 47.5 | 25.2 | 43.2 | 37.7 | 45.6 | 37.8 | 43.5 |
|  | Widowed/ separated | 12.4 | 4.9 | 12.1 | 4.3 | 14.3 | 6.6 | 12.4 | 8.5 | 15.3 | 4.9 | 13.0 | 5.3 | 13.7 | 2.6 | 10.5 | 2.3 |
| Educational level (%): None |  | 10.4 | 4.6 | 8.4 | 3.8 | 2.0 | 17.1 | 1.0 | 9.7 | 4.3 | 1.5 | 1.3 | 0.7 | 23.4 | 17.4 | 12.3 | 10.9 |
|  | Primary | 54.4 | 46.3 | 46.8 | 40.3 | 59.3 | 55.3 | 38.6 | 45.2 | 32.6 | 27.3 | 25.8 | 22.9 | 67.1 | 70.3 | 64.3 | 65.2 |
|  | Secondary | 35.1 | 49.1 | 44.8 | 55.9 | 38.7 | 27.6 | 60.4 | 45.1 | 63.1 | 71.2 | 72.9 | 76.4 | 9.6 | 12.3 | 23.4 | 24.0 |
| HIV prevalence (%) |  | 15.9 | 12.2 | 15.0 | 11.7 | 26.0 | 18.8 | 29.6 | 19.5 | 21.0 | 14.6 | 16.7 | 11.3 | 3.6 | 2.2 | 3.6 | 2.5 |
| Recent uptake of HIV testing (%) |  | 20.7 | 12.9 | 47.6 | 39.1 | 8.0 | 5.8 | 59.1 | 37.9 | 8.3 | 7.5 | 49.3 | 36.8 | 12.9 | 11.7 | 39.7 | 36.9 |
|  |  | Malawi |  |  |  | Tanzania |  |  |  | Kenya |  |  |  | Ethiopia |  |  |  |
| Survey Year |  | 2004 |  | 2015-16 |  | 2003-04 |  | 2011-12 |  | 2003 |  | 2008-09 |  | 2005 |  | 2016 |  |
| Gender |  | F | M | F | M | F | M | F | M | F | M | F | M | F | M | F | M |
| *Individual response rate (%) |  | 96 | 86 | 98 | 95 | 96 | 91 | 96 | 89 | 94 | 86 | 96 | 89 | 96 | 89 | 95 | 86 |
| Sample size (n) |  | 11,698 | 3,261 | 24,562 | 7,478 | 6,863 | 5,659 | 10,967 | 8,352 | 8,195 | 3,578 | 8,444 | 3,465 | 14,070 | 6,033 | 15,683 | 12,688 |
| Living in rural area (%) |  | 82.2 | 79.5 | 81.7 | 81.5 | 69.1 | 69.7 | 73.0 | 74.4 | 74.9 | 74.6 | 74.6 | 73.9 | 82.2 | 84.8 | 77.8 | 80.3 |
| Age (%): |  |  |  |  |  |  |  |  |  |  |  |  |  |  |  |  |  |
|  | 15-24 years | 45.0 | 37.9 | 42.4 | 43.1 | 41.8 | 41.8 | 39.2 | 42.3 | 43.3 | 43.0 | 41.2 | 40.6 | 41.3 | 39.8 | 39.2 | 35.1 |
|  | 25-34 years | 31.1 | 34.3 | 31.0 | 26 | 33.0 | 31.1 | 31.0 | 26.1 | 30.1 | 25.8 | 31.5 | 27.2 | 30.7 | 24.8 | 33.8 | 28.5 |
|  | ≥ 35 years | 23.9 | 27.8 | 26.5 | 30.8 | 25.2 | 27.1 | 29.8 | 31.5 | 26.6 | 31.2 | 27.3 | 32.2 | 28.0 | 35.5 | 27.0 | 36.4 |
| Family situation: Living in union |  | 71.1 | 63.8 | 65.7 | 58.1 | 63.6 | 53.1 | 63.0 | 53.0 | 60.0 | 50.8 | 58.4 | 51.4 | 64.4 | 56.8 | 65.2 | 58.9 |
|  | Single | 16.8 | 33.2 | 21.0 | 38.3 | 24.6 | 41.4 | 25.5 | 42.3 | 29.8 | 45.0 | 31.2 | 44.0 | 25.0 | 40.1 | 25.7 | 38.6 |
|  | Widowed/ separated | 12.1 | 3.0 | 13.3 | 3.5 | 11.9 | 5.5 | 11.5 | 4.7 | 10.2 | 4.2 | 10.4 | 4.6 | 10.6 | 3.2 | 9.1 | 2.5 |
| Educational level (%): None |  | 22.6 | 11.1 | 12.1 | 6.0 | 22.2 | 11.2 | 17.8 | 9.3 | 12.7 | 6.4 | 8.9 | 4.1 | 65.9 | 42.9 | 47.8 | 30.3 |
|  | Primary | 61.9 | 62.6 | 62.1 | 58.5 | 69.4 | 77.8 | 64.7 | 67.1 | 58.0 | 56.7 | 56.8 | 51.9 | 22.2 | 37.3 | 35.0 | 46.5 |
|  | Secondary | 15.5 | 26.3 | 25.8 | 35.5 | 8.4 | 11.0 | 17.5 | 23.6 | 29.3 | 36.9 | 34.3 | 44.1 | 11.9 | 19.8 | 17.2 | 23.2 |
| HIV prevalence (%) |  | 13.9 | 10.3 | 11.0 | 7.2 | 7.5 | 6.1 | 6.2 | 3.8 | 8.7 | 4.7 | 8.1 | 4.6 | 1.7 | 0.9 | 1.2 | 0.6 |
| Recent uptake of HIV testing (%) |  | 7.5 | 8.0 | 44.4 | 42.3 | 5.7 | 8.4 | 32.5 | 27.7 | 7.7 | 8.1 | 30.6 | 23.3 | 4.0 | 2.4 | 21.2 | 19.7 |

\*Based on each country's USAID DHS Final Report.

Participation rates varied between 90% and 99.5% among women and between 82% and 99.5% among men. Overall, data were collected among a total of 354,431 women and 183,353 men.

In the later surveys, in each country, the majority of the participants were living in rural areas (except for Liberia, Côte d’Ivoire and Cameroon) and living in union. Across surveys, HIV prevalence was lowest in Niger (0.7% in the earlier survey, 0.4% in the later survey) and highest in Lesotho (23.0% in in the earlier survey, 25.0% in the later survey) (Appendix 2).

### HIV testing

When considering males and females simultaneously (Figure 1 and Appendix 2), recent uptake of HIV testing was lowest in Niger (1.3%) in the earlier survey and in Guinea (5.0%) in the later survey. It was highest in Zambia (17.0%) in the earlier survey and in Lesotho (52.6%) in the later survey. For the earlier surveys (Table 1), women reported higher recent testing uptake than men in 8 out of 16 countries; while in the later surveys, women reported higher uptake than men in 15 out of 16 countries. With few exceptions (Lesotho 2014, women; Zimbabwe 2015, women), recent HIV testing was more frequently reported in urban than rural areas (Appendix 3).

**Figure 1:**
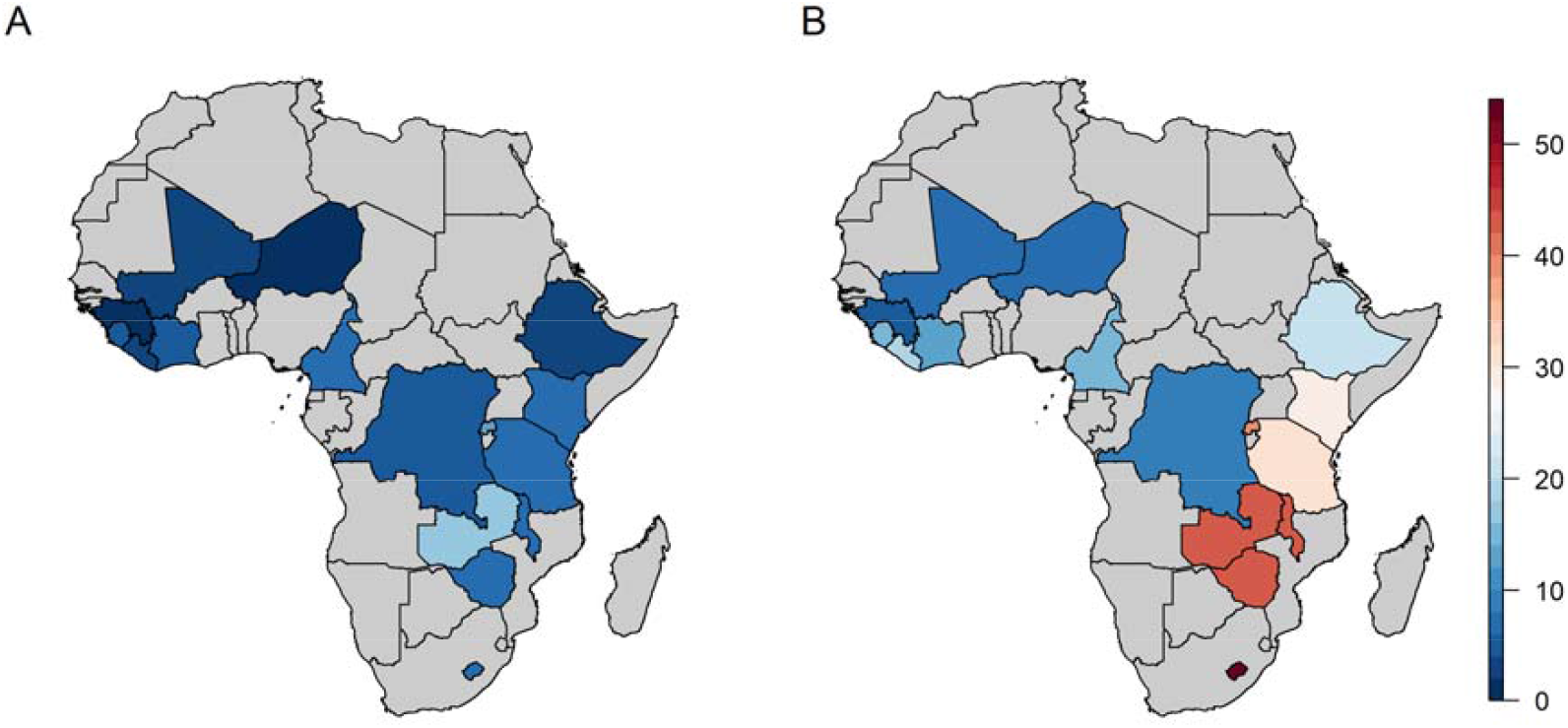
Percentage of recent (<12 months) uptake of HIV testing before (A) and after (B) 2008 in 16 sub-Saharan African countries. Percentage are those estimated from the later Demographic and Health Survey conducted before and after 2008 (see Table 1 for survey year).

Figure 2 presents, for each country, the proportions of recent HIV testing per survey round and gender among the richest and poorest wealth quintiles. Among both genders, we observed a pattern towards higher uptake of recent testing in the richest as compared to the poorest quintile for both survey rounds.

**Figure 2:**
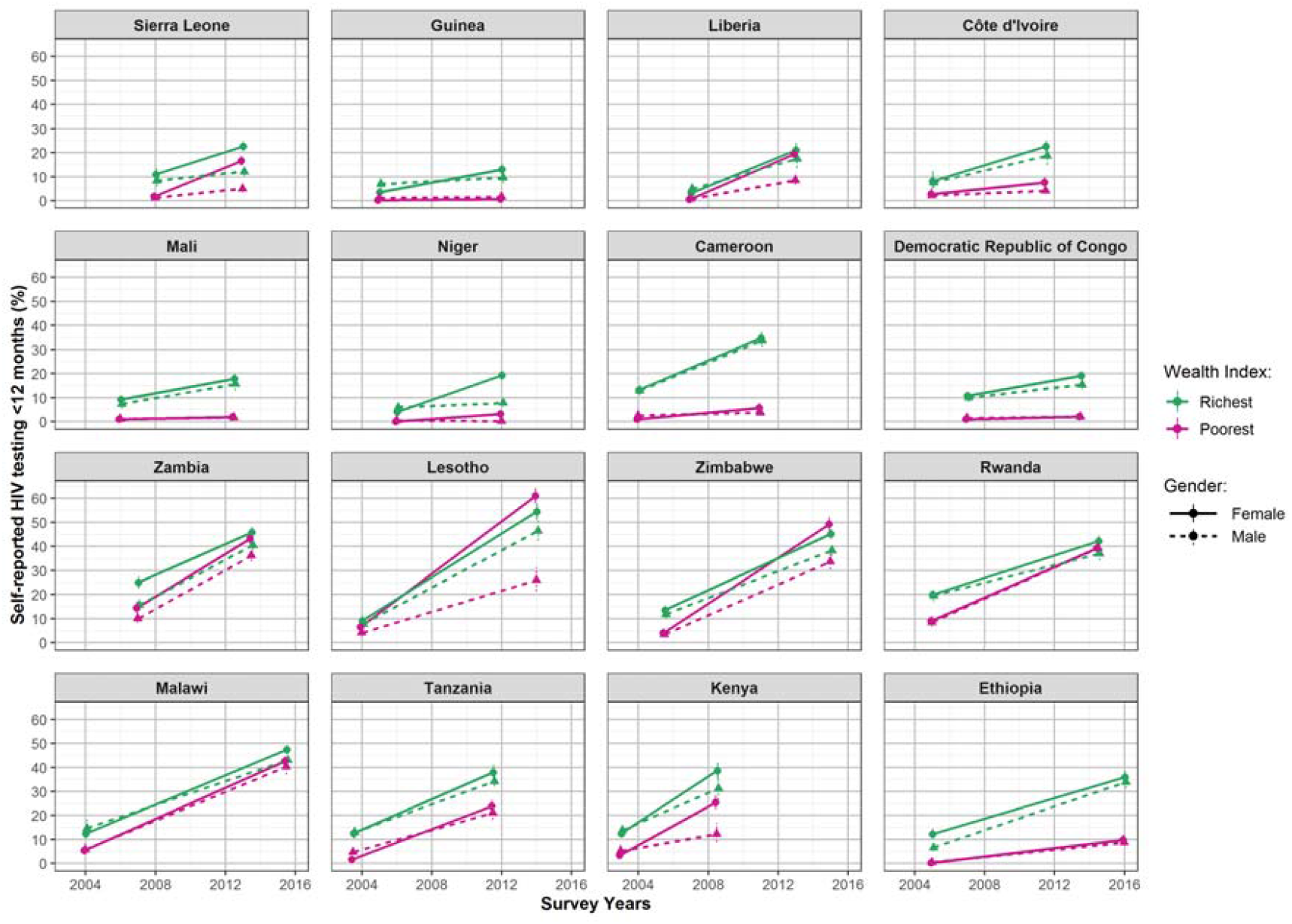
Gender-specific percentage of recent (<12 months) uptake of HIV testing among the richest and the poorest wealth quintiles between the pre- and post-treatment era in 16 sub-Saharan African countries. Countries are ordered west to east.

### Wealth-related inequalities in HIV testing

Gender-specific relative and absolute inequalities in recent HIV testing based on wealth distribution are presented in Tables 2 and 3. In the earlier surveys, pro-rich relative inequalities were observed in all 16 countries, for both men and women (all RII values >1). On average, before 2008, the wealthiest women were nearly 10 times more likely to report recent HIV test than the poorest (pooled RII 9.69, 95% CI 4.19-22.38). This ratio decreased to 2.69 after 2008 (95% CI 1.39-5.23; pooled standardized RII ratio 0.85.yr^-1^, 95% CI 0.80-0.90). However, in the later surveys, results were compatible with remaining inequalities in 13 out of 16 countries. This pattern was similar in men: we observed large pro-rich relative inequalities in earlier surveys, which decreased in the later survey (pooled standardized RII ratio 0.91.yr^-1^, 95% CI 0.86-0.96). However, inequalities remained in the later surveys in 14 out of 16 countries, with the richest men, on average, being 3.5 times more likely to report recent testing than the poorest (pooled RII 3.53, 95% CI 1.82-6.86). In the later surveys, relative inequalities were more marked in West and Central Africa as compared to Eastern and Southern Africa among both women and men (Wilcoxon rank-sum test: women, p-value = 0.007; men, p-value < 10^−3^).

**Table 2:** Relative and absolute wealth-related inequalities in self-reported recent (<12 months) HIV testing in women in 16 sub-Saharan African countries.

| Country |  | Relative inequalities |  |  | Absolute inequalities |  |  |
| --- | --- | --- | --- | --- | --- | --- | --- |
|  |  | Pre-2008 Survey RII | Post-2008 Survey RII | RII ratio (.yr<sup>-1</sup>) | Pre-2008 Survey SII | Post-2008 Survey SII | SII difference (.yr<sup>-1</sup>) |
| **Western and Central Africa** | **Sierra Leone** | 7.1 (4.5;11.4) | 1.4 (1.2;1.7) | 0.72 (0.66;0.80) | 0.11 (0.09;0.14) | 0.06 (0.03;0.09) | -0.011 (-0.019;-0.003) |
|  | **Guinea** | 135.6 (37.1;496.1) | 49.0 (29.2;82.1) | 0.86 (0.71;1.06) | 0.05 (0.04;0.07) | 0.17 (0.14;0.21) | 0.017 (0.012;0.022) |
|  | **Liberia** | 3.9 (2; 7.6) | 1.2 (1;1.4) | 0.82 (0.73;0.92) | 0.03 (0.01;0.04) | 0.04 (0;0.07) | 0.001 (-0.005;0.008) |
|  | **Côte d'Ivoire** | 5.4 (2.6;11) | 3.5 (2.8;4.4) | 0.94 (0.83;1.05) | 0.05 (0.03;0.08) | 0.19 (0.16;0.22) | 0.021 (0.014;0.027) |
|  | **Mali** | 34.3 (18.8;62.5) | 25.8 (17.5;38.1) | 0.96 (0.86;1.07) | 0.10 (0.08;0.13) | 0.23 (0.2;0.26) | 0.019 (0.013;0.025) |
|  | **Niger** | 58.2 (26.1;130.2) | 9.4 (7.1;12.5) | 0.74 (0.64;0.85) | 0.07 (0.05;0.09) | 0.23 (0.2;0.26) | 0.027 (0.021;0.033) |
|  | **Cameroon** | 29 (18.8;44.8) | 3.8 (3.1;4.6) | 0.75 (0.70;0.80) | 0.17 (0.15;0.2) | 0.15 (0.13;0.17) | -0.003 (-0.008;0.002) |
|  | **Congo DR** | 14.6 (9.4;22.9) | 12.9 (8.8;18.9) | 0.98 (0.90;1.07) | 0.14 (0.11;0.17) | 0.16 (0.14;0.19) | 0.003 (-0.003;0.009) |
| **Southern and Eastern Africa** | **Zambia** | 1.8 (1.4;2.2) | 1.1 (1;1.1) | 0.92 (0.89;0.96) | 0.12 (0.08;0.16) | 0.03 (-0.01;0.06) | -0.014 (-0.023;-0.006) |
|  | **Lesotho** | 1.4 (1.1;1.9) | 0.9 (0.8;0.9) | 0.95 (0.92;0.98) | 0.03 (0.01;0.05) | -0.1 (-0.14;-0.05) | -0.012 (-0.018;-0.007) |
|  | **Zimbabwe** | 5.6 (4.1;7.5) | 0.9 (0.9;1.0) | 0.83 (0.8;0.86) | 0.14 (0.11;0.17) | -0.04 (-0.08;0) | -0.019 (-0.024;-0.014) |
|  | **Rwanda** | 1.9 (1.6;2.4) | 1.1 (1.0;1.1) | 0.94 (0.92;0.96) | 0.09 (0.07;0.12) | 0.02 (-0.01;0.06) | -0.007 (-0.012;-0.003) |
|  | **Malawi** | 2.3 (1.7;3.1) | 1.0 (0.9;1.0) | 0.93 (0.91;0.95) | 0.06 (0.04;0.08) | -0.01 (-0.03;0.02) | -0.005 (-0.008;-0.003) |
|  | **Tanzania** | 9.2 (5.6;15.3) | 1.4 (1.2;1.5) | 0.79 (0.74;0.84) | 0.12 (0.09;0.15) | 0.10 (0.07;0.14) | -0.002 (-0.008;0.004) |
|  | **Kenya** | 5.5 (4; 7.5) | 1.6 (1.3;1.8) | 0.80 (0.75;0.85) | 0.13 (0.11;0.16) | 0.14 (0.09;0.18) | 0.001 (-0.009;0.01) |
|  | **Ethiopia** | 295.9 (170.9;512.6) | 4.6 (3.9;5.4) | 0.68 (0.65;0.72) | 0.38 (0.33;0.43) | 0.39 (0.34;0.43) | 0.001 (-0.005;0.007) |
| **Pooled estimate** |  | **9.79 (4.24;22.6)** | **2.77 (1.42;5.40)** | **0.85 (0.80;0.90)** | **0.11 (0.068;0.153)** | **0.111 (0.046;0.175)** | **0.001 (-0.006;0.008)** |
| **I<sup>2</sup>** |  | 97.75% | 99.00% | 94.19% | 95.72% | 97.68% | 95.36% |

RII: relative inequalities index, SII: slope index of inequalities. Average estimates and heterogeneity metrics ( $I^2$ ) are estimated from random-effect meta-analyses. RII-ratios and SII-differences are standardized on the number of years elapsed between both survey rounds. Countries are ordered west to east.

**Table 3:** Relative and absolute wealth-related inequalities in self-reported recent (<12 months) HIV testing in men in 16 sub-Saharan African countries.

| Country |  | Relative inequalities |  |  | Absolute inequalities |  |  |
| --- | --- | --- | --- | --- | --- | --- | --- |
|  |  | Pre-2008 Survey RII | Post-2008 Survey RII | RII ratio (.yr<sup>-1</sup>) | Pre-2008 Survey SII | Post-2008 Survey SII | SII difference (.yr<sup>-1</sup>) |
| **Western and Central Africa** | **Sierra Leone** | 7.5 (3.1; 18.0) | 2.8 (1.9; 4.2) | 0.82 (0.68; 1) | 0.09 (0.05; 0.13) | 0.09 (0.06; 0.13) | 0.001 (-0.01; 0.012) |
|  | **Guinea** | 32.3 (11; 95.2) | 10.3 (5.3; 20.3) | 0.85 (0.71; 1.02) | 0.10 (0.06; 0.14) | 0.12 (0.08; 0.16) | 0.002 (-0.006; 0.011) |
|  | **Liberia** | 10.7 (5.5; 20.9) | 3.4 (2.4; 4.8) | 0.83 (0.73; 0.93) | 0.06 (0.04; 0.08) | 0.15 (0.11; 0.2) | 0.015 (0.007; 0.023) |
|  | **Côte d'Ivoire** | 5.5 (2.5; 12.0) | 6.6 (4.3; 10) | 1.03 (0.9; 1.18) | 0.04 (0.02; 0.06) | 0.17 (0.13; 0.21) | 0.02 (0.013; 0.027) |
|  | **Mali** | 20.7 (8.7; 49.1) | 20.4 (11.5; 36.3) | 1.00 (0.85; 1.17) | 0.10 (0.06; 0.13) | 0.20 (0.15; 0.25) | 0.016 (0.007; 0.025) |
|  | **Niger** | 33.4 (12.6; 88.6) | 138.3 (47.9; 399.4) | 1.27 (1.00; 1.61) | 0.11 (0.07; 0.14) | 0.16 (0.12; 0.21) | 0.01 (0.001; 0.019) |
|  | **Cameroon** | 5.9 (4.1; 8.4) | 5.5 (4.5; 6.7) | 0.99 (0.93; 1.05) | 0.14 (0.11; 0.17) | 0.35 (0.31; 0.39) | 0.031 (0.024; 0.038) |
|  | **Congo DR** | 8.9 (5.2; 15.3) | 13.8 (8.7; 21.9) | 1.07 (0.96; 1.19) | 0.12 (0.09; 0.15) | 0.17 (0.13; 0.2) | 0.007 (0; 0.014) |
| **Southern and Eastern Africa** | **Zambia** | 1.9 (1.4; 2.5) | 1.2 (1.1; 1.3) | 0.93 (0.89; 0.97) | 0.09 (0.05; 0.12) | 0.06 (0.03; 0.09) | -0.004 (-0.012; 0.003) |
|  | **Lesotho** | 2.7 (1.6; 4.7) | 1.9 (1.6; 2.2) | 0.96 (0.91; 1.02) | 0.05 (0.02; 0.08) | 0.23 (0.17; 0.29) | 0.018 (0.011; 0.025) |
|  | **Zimbabwe** | 4.8 (3.4; 6.7) | 1.2 (1.1; 1.4) | 0.87 (0.83; 0.9) | 0.11 (0.09; 0.14) | 0.07 (0.03; 0.11) | -0.004 (-0.009; 0.001) |
|  | **Rwanda** | 3.2 (2.3; 4.3) | 0.9 (0.8; 1.1) | 0.88 (0.85; 0.91) | 0.14 (0.1; 0.18) | -0.02 (-0.07; 0.02) | -0.017 (-0.024; -0.011) |
|  | **Malawi** | 3.4 (2.0; 5.5) | 1.0 (0.9; 1.1) | 0.90 (0.86; 0.94) | 0.09 (0.05; 0.13) | 0.00 (-0.05; 0.04) | -0.008 (-0.013; -0.003) |
|  | **Tanzania** | 3.2 (2.3; 4.6) | 1.7 (1.4; 1.9) | 0.92 (0.88; 0.96) | 0.09 (0.06; 0.12) | 0.13 (0.09; 0.17) | 0.005 (-0.001; 0.011) |
|  | **Kenya** | 5.3 (3.4; 8.3) | 2.3 (1.7; 3.0) | 0.86 (0.78; 0.94) | 0.14 (0.1; 0.18) | 0.20 (0.13; 0.26) | 0.011 (-0.003; 0.024) |
|  | **Ethiopia** | 127.6 (56.3; 289.2) | 4.3 (3.7; 5.1) | 0.74 (0.68; 0.79) | 0.16 (0.13; 0.2) | 0.33 (0.3; 0.37) | 0.015 (0.011; 0.02) |
| **Pooled estimate** |  | **7.32 (4.09; 13.11)** | **3.55 (1.85; 6.81)** | **0.91 (0.86; 0.96)** | **0.101 (0.083; 0.119)** | **0.151 (0.096; 0.206)** | **0.007 (0.001; 0.014)** |
| **I<sup>2</sup>** |  | 91.41% | 98.27% | 80.72% | 79.47% | 96.02% | 92.69% |

RII: relative inequalities index, SII: slope index of inequalities. Average estimates and heterogeneity metrics ( $I^2$ ) are estimated from random-effect meta-analyses. RII-ratios and SII-differences are standardized on the number of years elapsed between both survey rounds. Countries are ordered west to east.

Important inequalities were also observed on the absolute scale among both men and women in all countries in the pre-2008 surveys (all SII values >0). There were however no identified changes in absolute inequalities in women (pooled difference in SII between pre- and post-2008 surveys 0.001.yr^-1^, 95% CI −0.006 - 0.008), and in the later surveys, on average, wealthier women still reported above 10 percentage points more than the poorest in recent HIV testing (pooled SII 0.112 95% CI 0.051-0.173). Among men, absolute inequalities increased on average between pre- and post-2008 surveys (pooled difference in SII between later and earlier surveys 0.007.yr^-1^, 95% CI 0.001-0.014).

When considering simultaneously relative and absolute inequalities, the countries that succeeded in reducing both relative and absolute inequalities after 2008, in both men and women, were Malawi, Rwanda, Zambia, and Zimbabwe.

### Education-related inequalities in HIV testing

Inequalities measured based on educational attainment exhibited similar patterns, with average relative inequalities decreasing in both genders, while absolute inequalities plateaued in women and increased in men (Appendix 4).

## Discussion

Using repeated cross-sectional population-based surveys, we provide a comprehensive assessment of inequalities in HIV testing uptake in sub-Saharan Africa, together with their time trends over the last 15 years. Uptake of HIV testing increased between pre- and post-2008 surveys in the 16 countries included in the analysis. HIV testing was more frequent in urban than rural areas nearly everywhere in the pre- and post-treatment era. Before 2008, testing uptake was roughly balanced between both genders; however, after 2008, women reported higher levels of recent testing than men in 15 out of 16 countries. Overall, we observed large inequalities in disfavor of the poor, both on the relative and absolute scales. Relative inequalities decreased over time among both genders, while absolute inequalities plateaued in women and increased in men. In the most recent surveys, important relative and absolute inequalities remained in a majority of countries.

We consistently observed increases over time in HIV testing uptake in both genders, as previously documented.^8^ Indeed, funding for HIV programs, including for HIV counselling and testing, dramatically increased during the era of treatment scale-up in sub-Saharan Africa.^21^ Concomitantly, the development and spread of new approaches for outreach and HIV testing allowed the intensification of HIV testing programs, notably with the expansion of provider-initiated HIV testing after 2007,^22^ and later on the development of community-based HIV testing.^2^ Despite encouraging increases in HIV testing in the past decades, efforts are still required in order to fulfill the target of 90% of people living with HIV knowing their status, especially in Western and Central Africa.^23^

We observed that after 2008, during the time of ART scale-up, women reported higher proportions of recent HIV testing than men nearly everywhere, a pattern that was not observed before 2008. Our analysis did not distinguish across HIV testing settings, but a global push on prevention of mother to child transmission, through providing provider-initiated routine testing and ART in antenatal clinics, may have largely contributed in the overall increase in testing among women.^22^ The lack of efforts to pursue the integration of HIV testing services into other relevant clinical settings may explain the gender-inequality disfavoring men with regards to HIV testing and treatment access and coverage, contributing to the blind spot surrounding men with regard to HIV prevention.^24^ Indeed, men are increasingly considered as a blind spot of the epidemic. Provider-initiated testing has been suggested to decrease socio-economic inequalities in HIV testing uptake.^6^ The higher levels of inequalities scales that we observed in men as compared to women in the recent surveys may thus also be linked to differential focus on opportunities of provider-initiated testing between genders.

The trends in inequalities we described were diverging whether considering relative or absolute measures of inequalities. Thus, one may draw different conclusions on the effect of HIV testing scale-up on inequalities if looking only at relative (decreasing inequalities) or absolute (plateaued or increasing inequalities) effect measures. Such a situation is actually quite frequent when studying health inequalities, and highlights the importance of using both absolute and relative effect measures when reporting inequalities.^15,25^ Relative inequalities tend to be larger at low overall levels of the considered outcome, while absolute inequalities tend to be larger at intermediate levels of the outcome.^26^ Thus, an increase in overall levels of HIV testing from low to intermediate levels between both surveys rounds is consistent with the inequalities trends described here, especially with increasing absolute inequalities observed in some Western and Central African counties. A corollary is that overall levels of HIV testing are to be considered when comparing different countries in terms of inequalities, especially when considering the absolute scale. Consider the case of women in the later surveys. Interpreting that Sierra Leone is doing better in terms of equity (SII=0.06) than Côte d’Ivoire (SII=0.19) is valid because the overall levels of recent testing are similar (17.6% and 15.4%, respectively). Conversely, it would be erroneous to judge that Sierra Leone is doing better in terms of equity than Kenya (SII=0.14) as the overall level of recent testing is nearly twice higher in the latter (30.6%).

Despite progress, at least on the relative scale, socio-economic inequalities remained substantial in the post-treatment era, especially in men. A better understanding of the sources of heterogeneity in the level of inequalities is required to better address them. The inequalities we observed do not only reflect differential access to HIV testing services in urban versus rural area. Indeed, socio-economic inequalities remain when accounting for this factor in multivariate analysis (Appendix 3). The burden of the HIV epidemic, which also drives the level of the response to it, seems to play a role in the pattern we observe. Indeed, countries with high HIV prevalence, such as Lesotho, Zimbabwe, Zambia and Malawi are also those where inequalities were less marked. Conversely, low-prevalence countries such as Niger, Ethiopia, Mali or Congo DR exhibited higher disparities in HIV testing uptake. Countries with high prevalence were also those prioritized for ambitious HIV testing programs, and HIV prevalence was found to be associated with HIV spending.^27^ This may suggest that low to moderate efforts to promote and offer HIV testing may perpetuate socio-economic inequalities, whereas larger efforts, even if not specifically targeted to low socio-economic positions, may decrease these inequalities.

Our analysis carries several limitations. Our results rely on a self-reported outcome. Assessing the validity of self-reports of HIV testing is challenging, notably because accuracy may differ depending on HIV status.^28,29^ As inequalities measurements rely on the quantification of an association, a differential accuracy in self-reporting between socio-economic groups could have conducted to biased results. To our knowledge, there is currently limited evidence regarding sensitivity and specificity of self-reported HIV testing depending on socio-economic conditions. However, evidence for other conditions such as cancer rather suggest that over-reporting of self-reported screening is higher for disadvantaged groups.^30^ If such a differential over-reporting also applies for HIV testing, this would have led to an under-estimation of the pro-rich inequalities we measured. It may also have contributed to the pro-poor inequalities observed in some countries (for women in Lesotho or Zimbabwe, for instance). Another limitation holds in the heterogeneity observed in the results of the meta-analysis, that prevented us to generalize our results beyond the subset of countries that we included in the analysis (Appendix 5). To our knowledge, this study is the first one to describe trends in socio-economic relative and absolute inequalities in the uptake of recent HIV testing across a large number of sub-Saharan African countries, covering a variety of regional and epidemiological contexts. Moreover, our analysis was based on large, representative surveys with high response rates, and the patterns of inequalities we describe were consistent across different dimensions used for their measure.

In conclusion, this study shows that overall increases in the uptake of HIV testing over the past decades hid differential progresses across socio-economics groups defined by gender, area of residence, wealth or education. Without specific focus on equity, HIV programs are unlikely to reach every part of the population, especially the poorest and the less-educated. Our results highlight the need to monitor and address socio-economic inequalities, among other forms of inequalities such as those based on gender and age in HIV programs in order to ensure an equitable distribution of their benefits.

## Data Availability

Data from the DHS surveys used in this article are publicly available for academic research (www.dhsprogram.com).

## Panel: Research in context

### Evidence before this study

Many studies documented socio-economic inequalities in the access to specific HIV prevention services, such as HIV testing, prevention of mother-to-child transmission or voluntary medical male circumcision, and to HIV treatment. We searched PubMed up to October 15th 2019 using the search (“inequality” OR “inequity” OR “equity”) AND (“HIV testing”) AND (“Africa”) without date or language restriction. The research returned 43 articles that we completed by screening the reference list of relevant articles. Most of the studies assessed socioeconomic inequalities in specific subgroups of the population, for instance pregnant women, or specifically focused on other forms of inequalities such as gender or age inequalities. Among studies focusing on wealth- or education-related inequalities, most focused on a single country (*ie* Uganda, South Africa, Côte d’Ivoire). They all identified wealth and/or education as predictors of HIV testing. One study (Cremin et al) assessed socioeconomic inequalities in the knowledge of one’s HIV status across 13 sub-Saharan countries in the pre-treatment era (before 2006) and documented a general trend to higher levels HIV status knowledge among wealthier and more educated individuals. Additionally, one grey-literature report (Staveteig et al) described the demographic characteristics associated with HIV testing in several sub-Saharan African countries, based on Demographic and Health Surveys conducted up to 2011. In gender-specific univariate analysis, uptake of HIV testing tended to increase monotonically with wealth. There were few exceptions, though, especially in countries with very high or very low overall levels of testing. Although socio-economic inequalities in HIV testing in sub-Saharan Africa have been documented in many studies, no pooled estimate are currently available. Moreover, whether these inequalities are reducing or worsening remains to be documented.

### Added value of the research

Based on standardized population-based surveys, we document the magnitude of wealth- and education-related inequalities in HIV testing together with their changes over time in 16 sub-Saharan African countries. We report both relative and absolute inequalities based on indicators that are widely used for quantification and comparison of socioeconomic gradients in health, and also calculate pooled estimates. In the most recent surveys, we observed a general trend toward inequalities disfavoring the poors and the less-educated. These socioeconomic inequalities were more sharped in men than women: on average wealthiest men were 3.5 time more likely to report recent testing than were the poorest; this ratio was 2.7 among women. Relative inequalities were deeper in Western- and Central-African countries as compared to Eastern- and Southern-African countries. When contrasting the pre- and post-2008 surveys, these inequalities decreased on the relative scale but plateaued among women and increased among men on the absolute scale.

### Implications of all the available evidence

After 2008, at the time of ART scale-up, socio-economic inequalities in recent HIV testing uptake remained substantial in many countries, despite reductions in relative inequalities. These results highlight the need to monitor progress in HIV testing uptake overall but also per sub-groups of population defined by socio-economic characteristics. A better understanding of the drivers of these inequalities is needed in order for current and future HIV testing policies to reach every part of the population, especially the poorest and the less-educated.

